# Research participants’ perception of ethical issues in stroke genomics and neurobiobanking research in Africa

**DOI:** 10.1101/2023.10.03.23296473

**Authors:** Ayodele Jegede, Olubukola Balogun, Olorunyomi Felix Olorunsogbon, Michelle Nichols, Joshua Akinyemi, Carolyn Jenkins, Mayowa Ogunronbi, Arti Singh, Reginald Obiako, Kolawole Wahab, Abiodun Bello, Albert Akpalu, Fred S. Sarfo, Lukman F. Owolabi, Babatunde Ojebuyi, Muyiwa Adigun, Dorcas Olujobi, Rabiu Musbahu, Musibau Titiloye, Ibukun Afolami, Benedict Calys-Tagoe, Ezinne Uvere, Ruth Laryea, Adekunle Fakunle, Osi Adeleye, Deborah Adesina, Nathaniel Mensah, Wisdom Oguike, Nathaniel Coleman, Sunday Adeniyi, Lanre Omotoso, Shadrack Asibey, Lois Melikam, Jibril Yusuf, Abdullateef Gbenga, Aliyu Mande, Muhammed Uthman, Rajesh N. Kalaria, Mayowa Owolabi, Bruce Ovbiagele, Oyedunni Arulogun, Rufus O. Akinyemi

**Author notes:** **Corresponding Author:**, (ROA).

## Abstract

**Background:** There is a growing interest in stroke genomics and neurobiobanking research in Africa. These raise several ethical issues, such as consent, re-use, data sharing, storage, and incidental result of biological samples. Despite the availability of ethical guidelines developed for research in Africa, there is paucity of information on how the research participants’ perspectives could guide the research community on ethical issues in stroke genomics and neurobiobanking research. To explore African research participants’ perspectives on these issues, a study was conducted at existing Stroke Investigation Research and Education Network (SIREN) sites in Nigeria and Ghana.

**Method:** Using an exploratory design, twenty-eight Focus Group Discussions (FGDs) sessions were conducted with stroke survivors (n=7), caregivers(n=7), stroke - free controls(n=7), and Community Advisory Board members(n=7). Data were collected using an interview guide. Interviews were conducted in English and indigenous languages of the community, audio recorded, and transcribed verbatim. Data were analyzed using NVivo (March, 2020) Software.

**Result:** Results revealed that stroke genomics and neurobiobanking research in Africa require researchers’ direct attention to ethical issues. Concerns were raised about understanding, disclosure and absence of coercion as components of true autonomous decision making in research participation. Participants argued that the risk and benefits attached to participation should be disclosed at the time of recruitment. Fears around data sharing were voiced as adherence to the principle of privacy and confidentiality were of paramount importance to participants. The preference was to receive the results of incidental findings with no stigma attached from society.

**Conclusion:** Research participants’ perspectives are a vital aspect of community engagement in stroke genomics and neurobiobanking research. Findings from this study suggest that research participants are interested in these fields of research in Africa if their concerns about ethical issues are appropriately addressed within the research framework.

## Introduction

As part of the efforts targeted towards combating the increasing prevalence and incidence of stroke and other cerebrovascular disorders (CVDs) in low- and middle-income countries (LMICs), especially in sub-Saharan Africa, genomics and neurobiobanking-based studies are on the increase. However, despite the scientific advancements, there exist challenges relating to ethical, legal, and social issues (ELSI). Hence, it is important to identify and develop strategies for addressing these challenges in the region.

For about a decade, the Human Heredity and Health in Africa (H3Africa) consortium has engaged in research on the African continent to generate data and samples that could be used to improve global health [1, 2]. In essence, they have developed numerous guiding documents on ethics, informed consent, governance, data sharing, and community engagement[3]. The African Neurobiobank for Precision Stroke Medicine (ELSI) project within the H3Africa consortium is a stroke genomic and neurobiobanking project across seven cities in Africa with the goal of identifying, examining, and developing novel approaches to address ELSI of biobanking and stroke genomic research in sub-Saharan Africa[4] Stroke genomics research raises several ethical questions and challenges [5], such as informed consent (who can give informed consent, type/nature of consent, what information should be contained in consent forms), sample ownership and management, sample sharing and storage, data storage and sharing, and role of regulatory committees are some of the ethical issues identified and broadly discussed in the literature[6].

Neurobiobanks are organized repositories for collection, storage, sharing and reuse of brain tissues, whole brain specimens and associated data for neurological research purposes [7]. These samples and data are used for a broad range of applied research activities in the development of tools tailored towards the prevention, diagnosis and treatment of diseases also known as precision medicine (PM). The samples and data are stored to be used for further research either in the countries of origin or other countries[7]. The field of neurobiobanking is growing in Africa with few studies already being conducted to understand the perspectives and willingness towards brain donation and brain banking among Africans[4, 8]. In addition, organized brain bank is being developed to accrue and store neural tissues for future research in neurology and psychiatry[9].

Genomics data deals with highly sensitive personal information which could significantly impact the lives of participants and their families. There is a need to consider ELSI in practice as there is still a dearth of literature about the consistent definition of key factors that constitute the ELSI variables in genomic research [10].

Though gaps exist in literature about what constitute and are defined as ethical criteria, most of these hinge on bioethical principles of beneficence, non-maleficence and autonomy, including those criteria that refer to human rights and dignity[10]. For reflection on ethical issues, empirical studies exploring research participants’ views and perceptions provide a good starting point to understanding stroke genomic and neurobiobanking research. Due to the complexity of the concepts and the vast cultural and geographical diversities in a multi-ethnic society, it is necessary to explore views from multicultural and diverse participants, including stroke survivors, caregivers, stroke free controls and Community Advisory Board (CAB) members.

In this study, we adopted Beauchamp’s and Childress’ four principles of beneficence, non-maleficence, autonomy and justice[11] for discussion of ethical issues in stroke genomic and neurobiobanking research. Autonomy is based on the principle of respect for persons to make their own choice or decision. The principle of beneficence is contained in the Hippocratic Oath which guides medical practice and is hinged on the principle of doing good or what will enhance patients’ interest. The ethical principle of non-maleficence requires that harm should be avoided to participants[12]

## Method

### Setting

The study was conducted in seven cities, five in Nigeria (Abeokuta, Ibadan, Ilorin, Kano and Zaria) and two in Ghana (Accra and Kumasi), as an integral part of an ongoing study [4].

### Study design

The study design was exploratory and employed Focus Group Discussions (FGDs) (S1 File) to investigate participants’ perception of ethical issues in stroke genomics and neurobiobanking research.

### Study population

The study population consisted of purposively selected stroke survivors, caregivers, stroke - free controls and Community Advisory Board (CAB) members between the ages of 18-85 involved in the SIREN study (Table 1) from seven sites in the established SIREN communities in Southern (Abeokuta and Ibadan), Central (Ilorin), and Northern (Kano and Zaria) Nigeria and Southern (Accra) and Central (Kumasi) Ghana[4]

**Table 1.** Focus group demographics.

| Study participants | Age range (years) | Males | Females | Total |
| --- | --- | --- | --- | --- |
| Stroke survivor | 22-80 | 27 | 22 | 49 |
| Stroke free controls | 22-65 | 31 | 18 | 49 |
| Stroke caregiver | 17-74 | 29 | 27 | 56 |
| Community Advisory Board | 27-82 | 45 | 14 | 59 |
| **Total** | **17-82** | **132** | **81** | **213** |

**Table 2:** Bioethical principles and themes.

| Bioethical principle | Themes |
| --- | --- |
| Autonomy | Understanding of research purpose |
|  | Disclosure of details of the research process |
|  | Coercion |
|  | Inducement |
| Beneficence | Individual Outcomes |
|  | Knowledge of treatment |
|  | Diagnosis of stroke disorder |
| Non-maleficence | Privacy and confidentiality |
|  | Data security |
|  | Stigmatization |

### Instrument

The FGD guide (S1 File) was developed from peer-reviewed literature relating to ELSI of neurobiobanking and stroke genomics, as well as lay literature in study communities/sites. The FGD guide explored issues around informed consent, data sharing, sample storage and reuse, feedback of research result and incidental findings, bio-right, governance, and regulation[4]

### Ethical consideration

All the seven sites obtained ethical approval from their institutional ethics review committee before the commencement of the study. University of Ibadan, Oyo State, Nigeria (UI/EC/18/0641); Federal Medical Centre, Abeokuta, Ogun State, Nigeria (FMCA/470/HREC/04/2019/01) Murtala Muhammad Specialist Hospital, Kano State, Nigeria (MOH/OFF/797/TI/1264); Ahmadu Bello University Teaching Hospital, Zaria, Kaduna State, Nigeria (ABUTHZ/HREC/E25/2018); University of Ilorin Teaching Hospital, Kwara State, Nigeria (UITH/CAT/189/19/007); University of Ghana College of Health Sciences, Accra, Ghana (EPRC/FEB/2021); Kwame Nkrumah University of Science and Technology, Kumasi, Ghana (CHRPE/AP/262/20). Written informed consent was obtained from each FGD participant. Participants were informed in detail about the study. Participation in this study was voluntary. The study participants were informed that they could withdraw from the study at any stage without any consequences to their benefiting from the outcome of the study, if any. To ensure confidentiality of study participants’ identity, numeric codes were allotted to each participant and only researchers had access to the data. Identifiers were removed from the transcripts.

### Data collection procedure, management, and analysis

A total of twenty- eight FGD sessions were conducted across all the seven sites between 26^th^ June 2019 and Feb 2020, and each session included between six and ten participants to ensure meaningful but actively guided discussions. CAB members were first briefed about the project during CAB meetings at each site and subsequently received updates at all CAB meetings. Other participants were approached and briefed by the study site coordinators about the study objectives. Participants were enrolled following informed consent. FGD sessions were held separately with each of the groups (stroke survivors, stroke caregivers, stroke-free control, and CAB members) in their respective sites. An interactive information sharing approach as a technique was adopted for this study, which involved participants providing information about stroke genomic and neurobiobanking research.

FGD sessions were conducted by a team comprising study sites coordinator and research assistants with experience in FGDs and qualitative research methods. Preceding the commencement of the FGDs, the research teams were trained on the protocol for data collection for this study. A pilot of the FGDs guide was carried out and revised before the actual data collection. The FGD sessions lasted on the average between 40-60 minutes.

All discussions were audio-recorded with the verbal consent of participants. The audio recordings were later transcribed verbatim and uploaded on REDCap, a secure data storage site. Transcripts were imported into NVivo (March 2020) software. The transcripts were reviewed and coded by three of the authors following the thematic framework for coding. Codes were generated in themes, inductively and deductively, both from literature and data [13]. The framework approach involving five stages were used which comprises:1) familiarization 2) identification of thematic framework 3) indexing 4) charting and 5) mapping and interpretation of data for data analysis.

## Results

A total of two hundred and thirteen persons (n =213) participated in the FGDs as presented in Table 1. The five themes identified from within the data were related to Beauchamp and Childress’ principles of biomedical ethics[11].

The analysis focused on exploring ethically relevant themes from research participants’ views, although the results are organized and presented according to Beauchamp and Childress’ principles of biomedical ethics[11]. Extracted text from participants’ perspectives were assigned to different themes and discussed iteratively.

### Autonomy

#### Stigmatization

Autonomy was explored regarding how participants viewed the informed consent process towards empowering decision making. Specifically, this pertains to how individuals voluntarily decide to participate in stroke genomic and neurobiobanking research. Majority of study participants indicated that there is the need to empower people with adequate information through the consent process by the researchers prior to participation to be able to express themselves voluntarily (Patient right). Participants expressed that for autonomous decision making (right to participate without coercion), an understanding of the purpose of the research, full disclosure of the research process and information about the study are important before consent.

#### Understanding of research purpose

Participants in this study indicated that a detailed explanation of the purpose of the research including the use of bio-samples should be expressly communicated before obtaining consent and participant understanding is crucial to decision making. Participants emphasized comprehensive understanding to prevent withdrawal. A respondent said:

> *What I understand…is before I join you in doing whatever you bring to me, you need to share information about what you want to do and make sure I understand it, get my feedback to say that I understand it. It is then I can agree if I can continue with you or not.* (Stroke - free control, Abeokuta)

A stroke caregiver respondent clarified that ‘*…the individual must be properly counseled about the purpose of the research, and she/he must understand it* (Stroke caregiver, Zaria).

CAB members across sites corroborated this view as indicated by a participant,

> “*What I know is that everything someone does, there is the need to brief her/him. Someone needs to be educated about it. If someone understands and accepts to do it…then she/he can sign for it”* (CAB member, Kumasi).

The central hallmark of the participants’ views is that participants’ empowerment will enable research participants make informed voluntary decisions. For them, once participants have adequate knowledge of the study it will be easy to make an informed decision.

#### Disclosure of details of research processes

Participants expressed the need to be provided with detailed information about the research processes including the risk and benefit associated with the use of samples obtained through the research.

> *Permission given with the full knowledge of the implications of their actions in terms of the side effects associated with the exact procedure that will be done on them, the products that will be obtained from them, how they will be obtained, and the consequences of it, if any…* (Stroke caregiver, Zaria)

This view was corroborated by the CAB participants saying that full disclosure of information about the research to be carried out for which participants would be recruited is important. According to a participant:

> *There must be full disclosure and the complete information must be provided regarding the purpose for this research* (CAB, Zaria).

This view was corroborated by the stroke survivor group according to one of them saying:

> *It is telling people all about the research - the good, the bad, and the ugly part of the research. Thereby participants are making an informed decision to be part of the studies*. (Stroke survivor, Zaria)

With respect to ethical concerns about neurobiobanking research, most respondents were in full support of sample usage, storage, and data sharing provided study participants are duly informed *So you have to inform the person that sample will be stored for future use because you cannot just take the sample and use it without informing the person* (Stroke Free control, Accra).

#### Coercion

Participants were of the view that consent should be sought without coercion. They argued that participants should have a good understanding of the reasons they should participate in the research. For instance, a participant said:

> *Consent that is given out of knowledge, for the fact that you know, not that you were compelled, or you are forced, or you are coerced…it means you have been told what it entails. (Abeokuta, CAB)*.

A stroke - free control participant emphasized the need for voluntariness (out of a free will) saying:

> *…that everything that you are doing (*participation) *is out of your own free will, you know nobody is forcing you to do it, every role you are to play, you are playing it freely and you are not doing it under duress. (Stroke - free control, Abeokuta)*.

#### Inducement

Participants emphasized that research participants should not be made to participate in research by giving or promising them what would make them lose their senses in making an irrational decision. For them, consent must be given freely without undue inducement. Adequate knowledge of the research activities was considered important for avoiding undue inducement. According to a participant:

> *You know what you are doing, you are not just giving the consent out of any inducement, it is done voluntarily, it is done by knowledge, it is done with full information at your disposal (CAB, Abeokuta)*.

#### Beneficence

With regards to the benefits of participating and sharing genetic information, participants across sites hold positive and inviting views towards stroke genomic research. The views are categorized into three themes: Individual outcomes, knowledge of treatment of stroke, and diagnosis of stroke. This will guide effective and informed participation in research.

#### Individual outcomes

Participants revealed that individual outcomes (benefits) as important ingredients in research participation. This view is rooted in the idea that individual outcomes of participating in research provide benefits to the community

> *…. You can share with others; we too can gain from their research. For example, they can do research we have not done, we can benefit from them. We too can-do research and they will benefit from us* (Stroke survivor, Ilorin).

However, participants echoed the need to be informed about the risk and potential benefits of the study before consenting to participate indicated that *…the benefits are outlined and if there are any risk too you are told…. (CAB, Accra)*.

This view was corroborated by the stroke - free control group in Ibadan saying:

> *‘The person must know about what he wants to get into and the benefits he will derive from it, the importance of the research, what he is going to gain from it before he can agree or disagree’. (Stroke -free control, Ibadan)*.Most of the participants corroborated the fact that participants benefit from research outcomes and individuals can show reciprocity by contributing to research for the benefit of humanity.

> *People have volunteered for research in the past that resulted in knowledge that is beneficial to my health so I would like to contribute to humanity* (Stroke Free Control, Ilorin).

Another potential benefit was expressed by a stroke survivor:

> *It is good, research helps to inform and make people and government aware of the appropriate medications for certain types of diseases. (Stroke survivor, Kano)*.

Participants’ perceptions of sample use for neurobiobanking research were positive as they posited the benefits to the society as stated by a stroke caregiver thus**…”***it is good because it makes diseases to reduce in our environment and anything that will reduce illnesses and diseases is good for our society. (Stroke caregiver, Abeokuta*). For the future advancement of genomic science ‘*It is good because you have something to fall back to when you want to do other genetic research* (Stroke survivor, Zaria). As well as benefits to humanity ‘*well, I think it a good thing I’m positively disposed to it, because the blood sample been kept is going to be used for research purposes that will benefit humanity so I’m positively disposed to it* (Stroke - free control, Abeokuta).

#### Discovery of treatment of stroke

Most of the participants across the study sites were in support of sharing data from stroke genomic research. They explained that the outcome of the research done with their samples will increase medical knowledge and practice in the treatment of stroke.

> *‘There is nothing wrong in that because it is the research some people do that gives us the knowledge of how to treat such diseases. So, if they do our own too and find out the causes of our disease and how the disease can be treated*…’. (Stroke survivor, Abeokuta).Similarly, a stroke caregiver corroborated the view saying:

> *‘When you share data…it will serve as a standard for other health officials to know how to go about it when someone has the disease and then you can care for people well*’. (Stroke caregiver, Accra).The Stroke survivor group agreed with this opinion as indicated by one of them who said:

> *…It will increase the knowledge of the researchers and the providers (*Stroke survivor, Kano).

Similarly, the Stroke caregiver group corroborated the views on the advantages of data sharing citing how it can improve the knowledge of healthcare providers and increase expertise in stroke genomic research. A respondent said:

> *‘Sharing of data and other findings concerning participants is very good, it is through that the health providers updates their knowledge and skills to treat patients’.* (Stroke caregiver, Kano).Participants also expressed the opinion that storage of bio-samples for research will enhance the advancement of knowledge about diseases and new innovations for treatments.

> *Storage of blood sample or fraction is very good, it is through those future investigations or research are conducted to know about the diseases and the new innovations on treatments and others (Stroke caregiver, Kano)*.

#### Diagnosis of stroke genetic disorders

Participants were unanimous that research could advance the process of establishing the diagnoses of stroke genetic disorders as well as finding new treatments. A participant said:

> *And the biobanking that you mentioned, I think it is very purposeful and if it is banked, the purpose of the banking is to ensure that you get the information so let’s say you have stroke or the sickness when your blood sample is banked, it helps to do more … So then you can also know the severity of the illness)* (Stroke survivor, Accra).

### Non-maleficence

There was a consensus among participants across the study sites about the use and sharing of data as well as the return of results. Several ethical concerns on conditions of use, data sharing, and return of result/incidental findings were emphasized by the participants.

#### Privacy and confidentiality

Participants’ echoed support for sharing of genetic information, if only identity will be protected.

> *...I don’t see anything wrong with that if it will not involve mentioning our names* (Stroke survivor Abeokuta).
>
> *‘Well, I don’t have any reservation about it and such information can be shared but about that as well will it be shared in a manner that will involve you saying this is my blood which contains something that I don’t want anyone to know and is it going to be shared in that manner? If it is going to be shared without my identity that shouldn’t be a problem.’* (CAB member, Accra).
>
> *I think if he would share it with another person, he has to seek my consent before he could share it with another person. If not, he is exposing my secret to someone else* (Stoke caregiver, Kumasi).

On the other hand, there was a participant who was not too concerned about the privacy and confidentiality of data,

> *I don’t have a problem with it. I gave it out so that it can be used to help others so I am okay with it being shared (Stroke survivor, Kumasi)*.

#### Data security

A relatively large number of participants expressed concerns about the security of data obtained from bio-sample data from participation in stroke genomic and neurobiobanking research

> *The issue of confidentiality is very important because if data is not properly managed there could be leakage which has legal implications that may result in suing ourselves* (Stroke survivor, Abeokuta).

#### Stigmatization

Most participants acknowledged the risk associated with stroke genomic and neurobiobanking research. A potential risk is stigmatization.

> *I think that stigmatization is also another factor, and we are in a society where people would always be stigmatized and given names based on their sickness and because of this people find it very difficult opening up, to give themselves up for a certain research work…* (Stroke survivor, Accra).

These views suggest that an individual’s health condition, especially when it is stigmatized, may influence response to persons experiencing or who had experienced one form of ill-health condition would want to participate in research on such condition or not.

## Discussion

The study explored participants’ perception of ethical issues in genomics and neurobiobanking using the Beauchamp and Childress (2001) ethical principles of autonomy, beneficence and non- maleficent. Data revealed similar sensitivities in perception from participants that understanding of research purpose, disclosure of details of the research process, coercion, inducement, individual outcomes, knowledge of treatment of stroke, diagnosis of stroke genetic disorders, privacy and confidentiality, data security, and stigmatization were critical ethical concerns associated with genomic and neurobiobanking research identified by participants across the study sites.

Participants were of the view that understanding and participation in stroke genomic and neurobiobanking research depends on how ethical issues and concerns are taken into consideration in the research process. Participants argued that the relationship between participants and researchers is shaped by community engagement strategy resulting in how well the potential participants understand the research process starting with the informed consent, use of the sample, and data sharing in an ethically safe approach. This finding agrees with other emerging body of knowledge from Africa on the collection, use, storage, and data sharing of biological samples [14–17]. Participants were of the view that adequate understanding of research during the consenting process is important for making well informed decision. This confirms the view expressed in an earlier study by [18] where it was argued that comprehension of information is an important ingredient to achieving free and informed consent decision making. Therefore, advancing research endeavours and promoting willingness especially in genomics and neurobiobanking requires innovative ways such as equipping researchers with adequate knowledge of the vocabularies in the field as well as translating them to the language of the participants for ease of comprehension. For example, we previously developed a mini dictionary showing definitions of vocabularies present in both our study instruments and the informed consent document [19]. This was translated to the main local languages in the different sites and each research assistant used this as guide when engaging the research participants in the language of their choice. This enhanced better understanding of the research focus and processes among the study participants.

While most participants viewed data sharing as a benefit, privacy and confidentiality of information were major ethical concerns raised by most of the participants. These views suggest that researchers have an obligation to accord respect to participants and allow them make an autonomous decision about their preference for sharing and re-use of their data. Participants were of the view that this should be agreed upon explicitly by both participants and researchers during the informed consent process. This finding agrees with views expressed by participants in a previous study with a similar theme [16, 20].

Coercion and inducement were viewed as twin ethical issues that need to be properly addressed so that participation in genomic and neurobiobanking research would be ethical. Free and informed consent, as the process of engaging potential participants, remains the solution to avoid coercion and inducement. Similarly, majority in our previous study opined that donation for genomic research should be a personal decision[21]. Due to the nature of genomic and neurobiobanking research, participation would be determined by how well participants are recruited and retained in the study. Coercion and inducement can be minimized by the researcher’s relationship with the potential participants. When trust is established with a particular researcher it can be extended to other researchers as earlier reported [22]. It is therefore imperative that researchers be transparent with participants so that they can gain their trust. They should ensure that the trust is sustained because once trust is lost it is difficult to recapture[23].

Closely associated with trust are issues of privacy and confidentiality. This is important because they determine, to a large extent, how participants would respond to requests for use, storage and sharing of data. All these bother on how much trust research participants have towards researchers. Research participants may use potential benefits as a condition for establishing trust in the researcher as earlier reported [22]. Therefore, providing unrealistic potential benefits may cause harm to the research process. However, researchers should endeavour to discuss both short- and long-term benefits of the research with the participants[24].

Participants also raised concern about the stigmatization of health conditions as an important issue to take into consideration during research engagement. This is because if the outcome of a diagnosis would result in a stigmatization, as is the case with stroke, participants may not want to participate. In this study participants indicated that stigmatization is a major risk associated with stroke genomic research. Despite large scale research on the challenges associated with stigmatized health conditions resulting in a large volume of scholarly publication in the literature, little attention has been devoted to addressing the ethical issues on stigmatized groups and health conditions[25]. Generally, a major ethical challenge identified with the stigmatized condition is the vulnerability of the person experiencing it[26]. This suggests the need for researching this area to understand how better to protect the vulnerable population.

## Conclusion

This study demonstrated that details of the research process, coercion, inducement, individual outcomes, knowledge of treatment of stroke, diagnosis of stroke genetic disorders, privacy and confidentiality, data security and stigmatization were ethical issues associated with genomic and neurobiobanking research in this cohort of West African research participants. Participants were of the view that understanding and disclosure of detailed information about the research during the consenting process would enable potential research participants make free and informed choices. Also, adequate knowledge of the research process through disclosure of information about the research in the informed consent process would reduce coercion and inducement as well as promote trustworthiness between participants and researchers. Participants were positive towards participation in neurobiobanking research but have specific ethical concerns that should be addressed within local contexts. Finally, the study showed that the ethical issues around stigmatizing health conditions are a major hindrance to a willingness to participate in research by persons experiencing such stigmatizing health conditions.

Researchers should take into consideration the ethical concerns of stroke genomic and neurobiobanking research participants in order to gain their trust and sustain their participation. The researcher should also ensure that participants are fully informed about what they need to know about the use, re-use, and sharing of their data and/or biological samples.

## Data Availability

All relevant data are within the manuscript and its Supporting Information files.

## Supplementary information

S1 File. Focus group discussion guide.

## References

1. Mulder N, Abimiku A, Adebamowo SN, de Vries J, Matimba A, Olowoyo P, et al. H3Africa: current perspectives. Pharmgenomics Pers Med. 2018;11:59–66.

2. Rotimi C, Abayomi A, Abimiku A, Adabayeri VM, Adebamowo C, Adebiyi E, et al. Research capacity. Enabling the genomic revolution in Africa. Science. 2014;344(6190):1346-8.

3. H3Africa Ethics Working Group. Ethics and governance framework for best practice in genomic research and biobanking in Africa. 2018.

4. Akinyemi RO, Jenkins C, Nichols M, Singh A, Wahab K, Akpalu A, et al. Unraveling the Ethical, Legal, and Social Implications of Neurobiobanking and Stroke Genomic Research in Africa: A Study Protocol of the African Neurobiobank for Precision Stroke Medicine ELSI Project. Int J Qual Methods. 2020;19.

5. De Vries J, Bull SJ, Doumbo O, Ibrahim M, Mercereau-Puijalon O, Kwiatkowski D, et al. Ethical issues in human genomics research in developing countries. BMC medical ethics. 2011;12:1–10.

6. Joly Y, Dalpé G, So D, Birko S. Fair shares and sharing fairly: a survey of public views on open science, informed consent and participatory research in biobanking. PloS one. 2015;10(7):e0129893.

7. Freund M, Taylor A, Ng C, Little AR. The NIH NeuroBioBank: creating opportunities for human brain research. Handbook of Clinical Neurology. 150: Elsevier; 2018. p. 41–8.

8. Akinyemi R, Ojagbemi A, Akinyemi J, Salami A, Olopade F, Farombi T, et al. Gender differential in inclination to donate brain for research among Nigerians: the IBADAN Brain Bank Project. Cell Tissue Bank. 2019;20(2):297–306.

9. Akinyemi RO, Salami A, Akinyemi J, Ojagbemi A, Olopade F, Coker M, et al. Brain banking in low and middle-income countries: Raison D’etre for the Ibadan Brain Ageing, Dementia And Neurodegeneration (IBADAN) Brain Bank Project. Brain Res Bull. 2019;145:136–41.

10. Ascencio-Carbajal T, Saruwatari-Zavala G, Navarro-Garcia F, Frixione E. Genetic/genomic testing: defining the parameters for ethical, legal and social implications (ELSI). BMC Medical Ethics. 2021;22(1):1–15.

11. Beauchamp TL, Childress JF. Principles of biomedical ethics: Oxford University Press, USA; 2001.

12. Iserson KV. Principles of biomedical ethics. Emergency Medicine Clinics. 1999;17(2):283–306.

13. Nowell LS, Norris JM, White DE, Moules NJ. Thematic analysis: Striving to meet the trustworthiness criteria. International journal of qualitative methods. 2017;16(1):1609406917733847.

14. Tindana P, Molyneux CS, Bull S, Parker M. Ethical issues in the export, storage and reuse of human biological samples in biomedical research: perspectives of key stakeholders in Ghana and Kenya. BMC Medical Ethics. 2014;15(1):1–11.

15. Bukini D, Mbekenga C, Nkya S, Purvis L, McCurdy S, Parker M, et al. A qualitative study on aspects of consent for genomic research in communities with low literacy. BMC medical ethics. 2020;21(1):1–7.

16. Matandika L, Ngóngóla RT, Mita K, Manda-Taylor L, Gooding K, Mwale D, et al. A qualitative study exploring stakeholder perspectives on the use of biological samples for future unspecified research in Malawi. BMC medical ethics. 2020;21(1):1–10.

17. Ochieng J, Kwagala B, Barugahare J, Mwaka E, Ekusai-Sebatta D, Ali J, et al. Perspectives and ethical considerations for return of genetics and genomics research results: a qualitative study of genomics researchers in Uganda. BMC medical ethics. 2021;22:1–9.

18. Jegede AS. Understanding informed consent for participation in international health research. Developing world bioethics. 2009;9(2):81–7.

19. Uvere EO, Nichols M, Ojebuyi BR, Isah SY, Calys-Tagoe B, Jenkins C, et al. Capacity- Building for Stroke Genomic Research Data Collection: The African Neurobiobank Ethical, Legal, and Social Implications Project Experience. Biopreserv Biobank. 2023;21(2):158–65.

20. Erdmann A, Rehmann-Sutter C, Bozzaro C. Patients’ and professionals’ views related to ethical issues in precision medicine: a mixed research synthesis. BMC medical ethics. 2021;22(1):116.

21. Singh A, Arulogun O, Akinyemi J, Nichols M, Calys-Tagoe B, Ojebuyi B, et al. Biological sample donation and informed consent for neurobiobanking: Evidence from a community survey in Ghana and Nigeria. PLoS One. 2022;17(8):e0267705.

22. Berrios C, James CA, Raraigh K, Bollinger J, Murray B, Tichnell C, et al. Enrolling genomics research participants through a clinical setting: the impact of existing clinical relationships on informed consent and expectations for return of research results. Journal of genetic counseling. 2018;27:263–73.

23. Jegede AS. What led to the Nigerian boycott of the polio vaccination campaign? PLoS medicine. 2007;4(3):e73.

24. Bedeker A, Nichols M, Allie T, Tamuhla T, van Heusden P, Olorunsogbon O, et al. A framework for the promotion of ethical benefit sharing in health research. BMJ Glob Health. 2022;7(2).

25. Millum J, Campbell M, Luna F, Malekzadeh A, Karim QA. Ethical challenges in global health-related stigma research. BMC medicine. 2019;17:1–9.

26. Tekola F, Bull S, Farsides B, Newport MJ, Adeyemo A, Rotimi CN, et al. Impact of social stigma on the process of obtaining informed consent for genetic research on podoconiosis: a qualitative study. BMC medical ethics. 2009;10:1–10.

